# Characterizing the Modern Light Environment and its Influence on Circadian Rhythms

**DOI:** 10.1101/2020.10.21.20214676

**Authors:** Dennis Khodasevich, Susan Tsui, Darwin Keung, Debra J. Skene, Victoria Revell, Micaela E. Martinez

## Abstract

Humans have largely supplanted natural light cycles with a variety of artificial light sources and schedules misaligned with day-night cycles. Circadian disruption has been linked to a number of disease processes, but the extent of circadian disruption among the population is unknown. We measured light exposure and wrist temperature among residents of an urban area for a full week during each of the four seasons, as well as light illuminance in nearby outdoor locations. Daily light exposure was significantly lower for individuals, compared to outdoor light sensors, for all four seasons. There was also little seasonal variation in the realized photoperiod experienced by individuals, with the only significant difference between winter and summer. We tested the hypothesis that differential light exposure impacts circadian phase timing, detected via the wrist temperature rhythm. To determine the influence of light exposure on circadian rhythms, we modeled the impact of morning, afternoon, and nighttime light exposure on the timing of the midline-estimating statistic of rhythm (MESOR). We found that morning light exposure and nighttime light exposure had a significant but opposing impact on MESOR timing. Our results demonstrate that nighttime light can shift/alter circadian rhythms to delay the morning transition from nighttime to daytime physiology, while morning light can lead to earlier onset. Our results demonstrate that circadian shifts and disruptions may be a more regular occurrence in the general population than is currently recognized.

**Significance Statement:** Disruption of circadian rhythms has been linked to various diseases, but the prevalence of circadian disruption among the general population is unknown. Light plays a pivotal role in entraining circadian rhythms to the 24-hour day. Humans have largely supplanted natural light cycles with electrical lighting and through time spent indoors. We have shown that individuals experience a disconnect from natural light cycles, with low daytime light exposure, high levels of light-at-night, and minimal seasonal variation in light exposure. We identified measurable changes in the timing of wrist temperature rhythms as a function of differential light exposure during the morning and nighttime hours. Our findings suggest that circadian shifts, and potentially disruption, may be common in the general population.

## Introduction

Circadian rhythms underlie many foundational biological processes across all corners of life, ranging from prokaryotes to humans [1]. Life evolved under predictable day-night cycles. Structuring certain biological processes into 24-hour cycles allowed organisms to maximize their fitness by synchronizing their internal biology with the external environment [2]. In mammals, nearly all aspects of physiology operate under some level of circadian control, resulting in orchestration of physiological conditions to appropriately match 24-hour cycles in the environment [3]. Well-documented circadian rhythms in mammals include: direct trafficking of various immune cells among the blood and organs [4], generating daily variation in gene transcription [5], controlling rhythms in the rate of protein translation [6], and altering functional responses to infection/vaccination [7,8]. Similarly, rhythms in melatonin, DNA damage, lipid peroxidation, and protein oxidation suggest there is circadian control involved in the response to oxidative stresses [9].

In addition to circadian rhythms, mammals display endogenous seasonal (i.e. circannual) rhythms in physiology [10]. Unlike the master circadian clock of mammals, located in the hypothalamic suprachiasmatic nuclei, which has been extensively studied, the circannual clock has yet to be revealed in great detail. Though its molecular architecture is unknown, the circannual clock of many mammal and bird species entrains to photoperiod (day length) [11] and may regulate changes in immunity and health [12]. The working conceptual model for seasonal rhythm generation is that the master circadian clock, which entrains itself to light, is modified seasonally as the duration of daylight changes with the seasons. Seasonal modulation of circadian rhythms could thus generate circannual rhythms [11,13].

Although the daily and seasonal light cycles that life evolved under continue to exist, humans have largely supplanted these natural light cycles with increased time spent indoors and new light cycles built around a variety of electrical light sources. Indoor lighting places humans in an illuminance setting that would not be experienced in nature. Electrical light experienced after sunset, termed light-at-night (LAN), can introduce high levels of light exposure at times that would normally be characterized by exceedingly low light exposure due to the lack of direct sunlight. Similarly, outdoor lighting and light spilling from buildings causes brightness many times above the lux of moonlight within urban areas and in the skies, termed light pollution. Unlike other exposures, LAN does not cause direct toxicity to the body, but instead causes perturbations to the circadian system with downstream physiological consequences [14]. The evolved use of light for rhythm entrainment can have pathological consequences in the presence of artificial light, such as elevated breast cancer risk due to light pollution and light at night [15]. A growing body of evidence suggests that chronic circadian disruption can contribute to the development of various diseases, including asthma, cancer, metabolic syndrome, and cardiovascular disease [16,17,18].

People live with their own unique realized light cycles, made up of a combination of natural sunlight, ambient light pollution, and indoor artificial lighting. The extent to which daily variations in realized light cycles disrupt circadian physiology in the real-world is poorly understood. In this study we set out to characterize the realized light cycles (RLC) of people in their normal environments, compared to outdoor light cycles, and identify any associations between variations in light exposure and variations in circadian physiology. Body temperature is under circadian control and has been used for many decades to monitor the circadian clock [19]. Due to the ease of measuring body temperature using wearable devices, we used wrist temperature as a non-invasive readout of the circadian system [20]. Our overall aim was to test three hypotheses. First, *people dim out their days* through time spent indoors and *light up their nights* through the use of electric light. Second, we hypothesize people experience relatively uniform light exposure throughout the year instead of the natural seasonal light cycle. Lastly, differential light exposure experienced during a normal routine can lead to shifts in circadian physiology, as detected in changes to the timing of wrist temperature rhythms.

## Results

### Light Exposure Around the Clock & Through the Seasons

Time series of light measured from our outdoor sensors was highly regular, relative to individual exposure, and tightly linked to local sunrise and sunset times (Figure 1a-b). There were minimal nonzero lux readings occurring outdoors after sunset, despite the relatively high amount of light pollution expected in New York City, our primary sample site. Given the lower limit of detection of the light sensors, the illuminance of the outdoor light pollution in the study area measured less than 10 lux. Due to the tight link with sunrise/sunset, seasonal changes in photoperiod were clearly observable from the outdoor sensors (Figure 1a). In contrast to outdoor light, individual light cycles exhibit a high degree of variation within and between individuals. Study participant light exposure did not closely align with sunrise and sunset times (Figure 1b). In particular, participants experienced high levels of nighttime light exposure and a high degree of variability in the degree of nighttime light exposure. Relative to outdoor light, individual light exposure time series featured a high number of days with low levels of light exposure. For instance, compared to a shaded outdoor area, which regularly reached maximum daily lux values of 10^3^-10^5^ lux, individual light exposure rarely exceeded 10^3^ lux and daily patterns were highly erratic (Figure 1b). Lastly, many individuals exhibited low levels of light exposure throughout most of their observation weeks, with one or two days of high intensity light exposure more closely resembling outdoor light readings, typically occurring on weekends.

**Figure 1:**
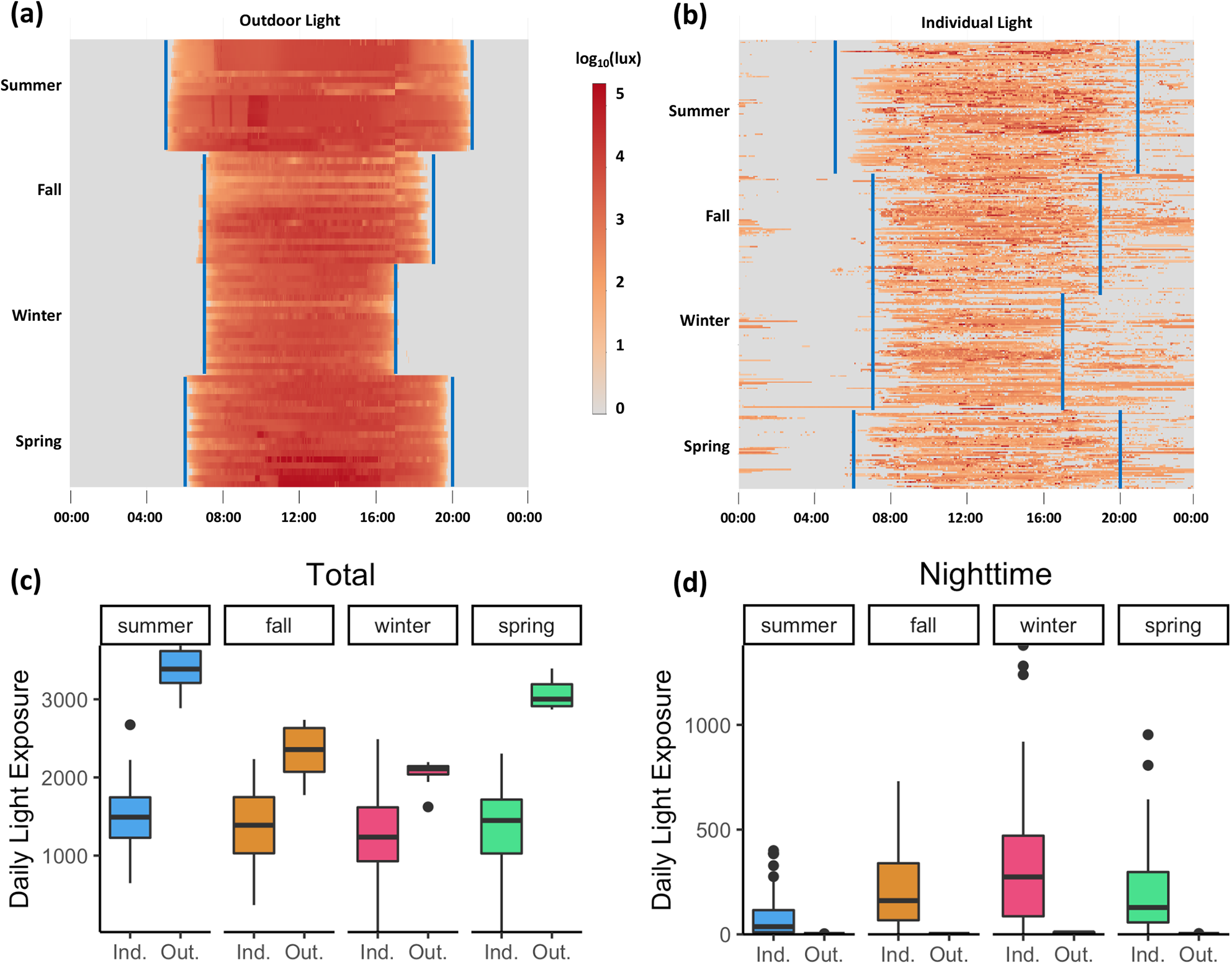
Light Data Characterization. Daily light exposure time series across all four seasons with approximate sunrise and sunset times shown in blue from (a) outdoor sensors and (b) individual light exposure. Each row contains a full 24-hour period of readings from a single light sensor, with rows grouped together by light sensor and by season. Individual data consists of lux readings taken at 5-minute intervals over up to 7 days from study participants. Outdoor data consists of lux readings taken at 3-minute intervals over 9 days from sensors located in upper Manhattan. (c) Total daily light exposure, measured as the area-under-the-curve for the log10lux timeseries, comparisons of individual participant data (Ind.) and outdoor data (Out.) across the four seasons. (d) Nighttime light exposure (Sunset – 04:00) comparisons of individual participant data (Ind.) and outdoor data (Out.) across the four seasons.

As for seasonal light exposure, individuals had relatively similar total daily light exposure from season-to-season, and the amount of light experienced was lower relative to outdoor light for all four seasons (Figure 1c). Differences in total daily light exposure measurements (the area under the curve of the log_10_lux time series) are highlighted in Table S1. Total daily outdoor light exhibited a seasonal pattern with light highest in the summer and lowest in the winter measured by outdoor sensors. However, total daily light exposure experienced by study participants exhibited no discernible seasonal pattern (Figure 1c; Table S1). As for nighttime light, most individuals experienced some nighttime light, even though little-to-no nighttime light was detected from the outdoor sensors (Figure 1d). Thus, we infer that nighttime light exposure came from the use of indoor lighting as opposed to outdoor light pollution exposure. When we partitioned the 24-hour cycle into morning, afternoon, evening, and late night we found that individuals experienced the most variation in light exposure late night (relative standard error, RSE = 11.83), while the most consistent light exposure was in the afternoon (RSE = 2.06).

### The Effect of Light-at-Night and Morning Light on Circadian Physiology

Individuals exhibited a large degree of variation in daily wrist temperature but followed the same general trend of reaching a maximum wrist temperature in the late night/early morning and falling to a minimum wrist temperature in the afternoon (Figure 2a). From this we infer that the nighttime physiological state consists of warm peripheral temperature and the daytime state consists of cool peripheral temperature. The transition from nighttime to daytime physiology tended to occur in the hours around sunrise, with seasonal variation in how closely aligned the transition was to sunrise (Figure 2a). The transition to nighttime physiology was not clearly aligned with sunset and this transition time tended to be noisier among individuals and between seasons (Figure 2a). Individual daily temperature trends were relatively noisy due to periods with missing data and the presence of high frequency variation in temperature within the overall 24-hour trend. The most common times for daily maximum temperature were 00:00 and 01:00 (Figure 2b).

**Figure 2:**
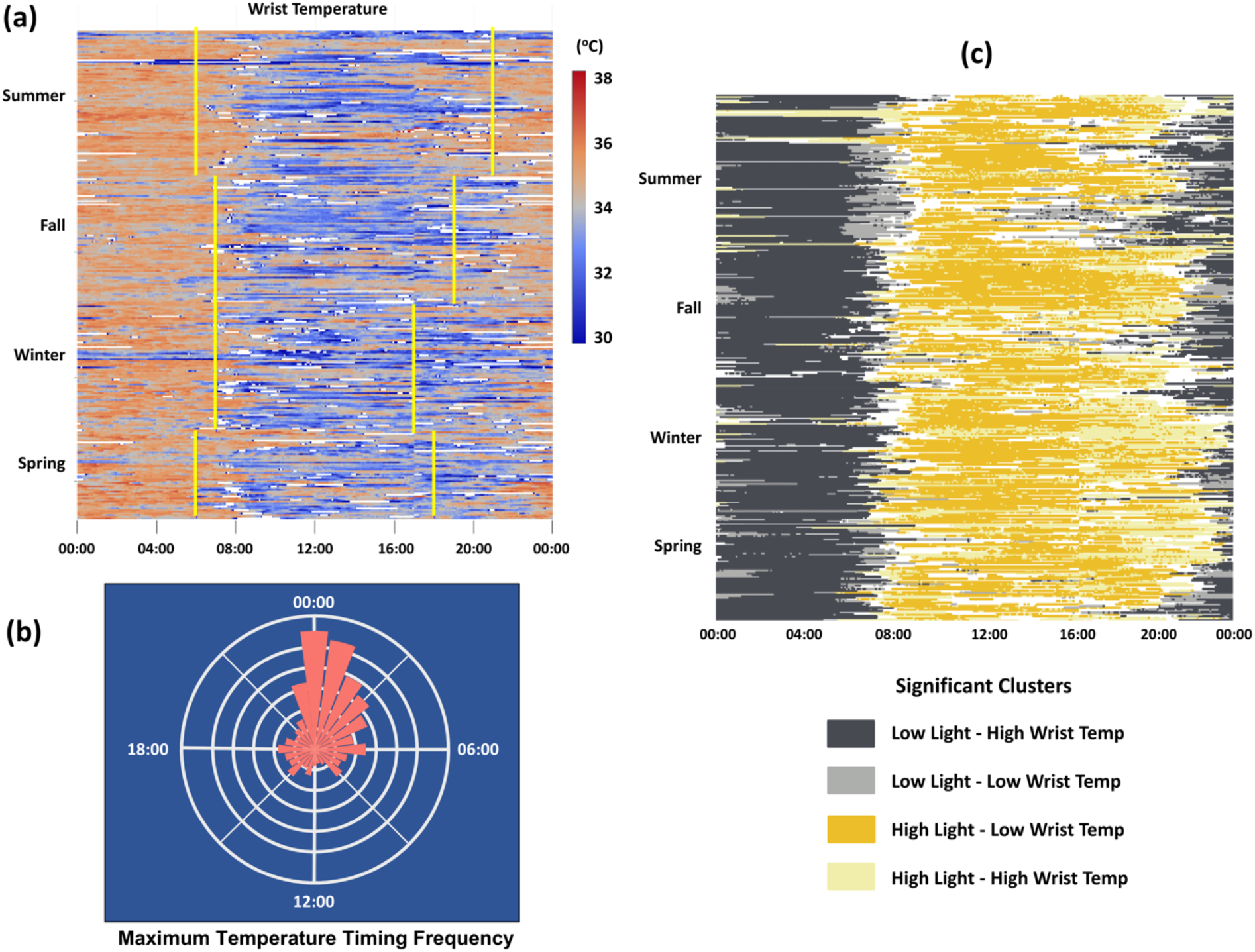
Wrist Temperature Characterization. (a) Daily wrist temperature time series across all four seasons with approximate sunrise and sunset times shown in yellow. Each row contains a full 24-hour period of readings from a single individual, with rows grouped together by individual and by season. Individual data consists of lux readings taken at 5-minute intervals over up to 7 days from study participants. Times with wrist temperatures outside of the range of normal human wrist temperature (< 29.5°C or > 38.5°C) appear as white cells. (b) Relative frequency of daily maximum temperature timing, based on hourly averages. (c) Local bivariate Moran’s I cluster analysis of individual light exposure and wrist temperature trend data. Significant clusters are shown in their corresponding colors, with non-significant areas shown in white.

The cluster analysis identified significant clusters shared among the light exposure and wrist temperature matrices (Figure 2c). The two most frequent and biologically relevant clusters were low light/high wrist temperature clusters and high light/low wrist temperature clusters. Low light exposure and high wrist temperature was indicative of nighttime physiology, while high light exposure and low wrist temperature was indicative of daytime physiology. The transition from nighttime physiology to daytime physiology typically occurred between 06:00 and 08:00, while the transition from daytime physiology to nighttime physiology was more variable.

The timing of the morning MESOR, which we define as the time point between peak nighttime wrist temperature and trough daytime wrist temperature, was used as a biological readout of the circadian phase. We used this readout specifically because it has the potential to be influenced by both nighttime light and daytime light. Furthermore, based on our cluster analysis, the morning MESOR coincides with the transition from nighttime physiology to daytime physiology (Figure 2c). The mean timing of the morning MESOR was at 08:40 with a large standard deviation of approximately 2.8 hours. Our linear mixed model tested the effect of nighttime, morning, and afternoon light on MESOR timing. There was a significant effect of nighttime light which caused the MESOR to occur later, as well as a significant effect of morning light, which shifted the MESOR later, as well as no significant effect of afternoon light (Table 1). Baseline MESOR timing varied substantially among participants, ranging from 07:00 to 11:30. The effect size of morning and nighttime light exposure were relatively similar in magnitude, suggesting they may have equal but opposing effects on when the body transitions from nighttime to daytime physiology.

**Table 1:**
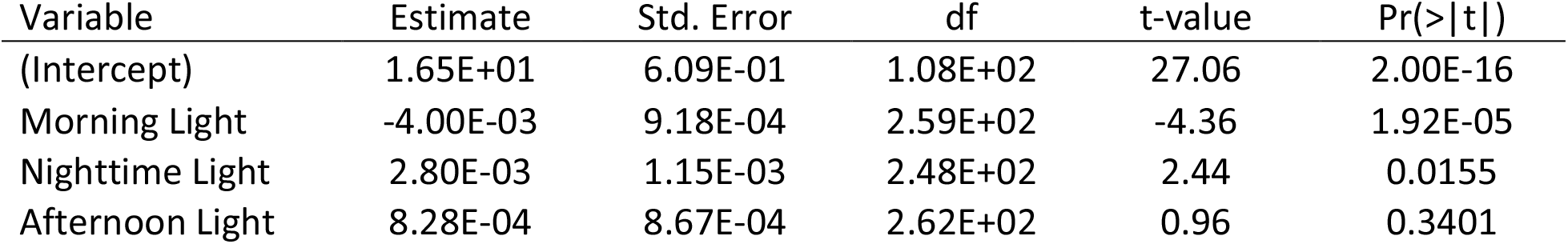
Fixed Effects Outputs from the Linear Mixed Model. Linear mixed model fit by REML, t-tests use Satterthwaite’s method. Accompanying p-values for the fixed effects are under the “Pr(>|t|”) column. Random effects allow for y-intercept to vary between individuals. All input variables had a variance inflation factor below 1.47, suggesting no multicollinearity among the variables.

Overall, more morning light and less nighttime light exposure was associated with earlier MESOR timing, while less morning light and more nighttime light exposure was associated with later MESOR timing (Figure 3). There are multiple ways in which circadian rhythms can be modulated to generate the observed shift in MESOR timing found by our model. One potential method is through an overall phase shift, in which the entire daily wrist temperature cycle is moved earlier or later due to the timing of light exposure. Specifically, morning light exposure may shift the entire temperature rhythm, generating an earlier morning MESOR timing (Figure S4a), while nighttime light exposure shifts the MESOR timing later (Figure S4b). Another potential process is through an alteration of the cycle/rhythm shape, in which the normal daily wrist temperature rhythm is temporarily distorted by light exposure. For instance, morning light exposure may lead to a faster decline in wrist temperature (Figure S4c), as opposed to a phase shift. Similarly, nighttime light may lead to a delayed rise in wrist temperature and/or other distortions of the rhythm (Figure S4d).

**Figure 3:**
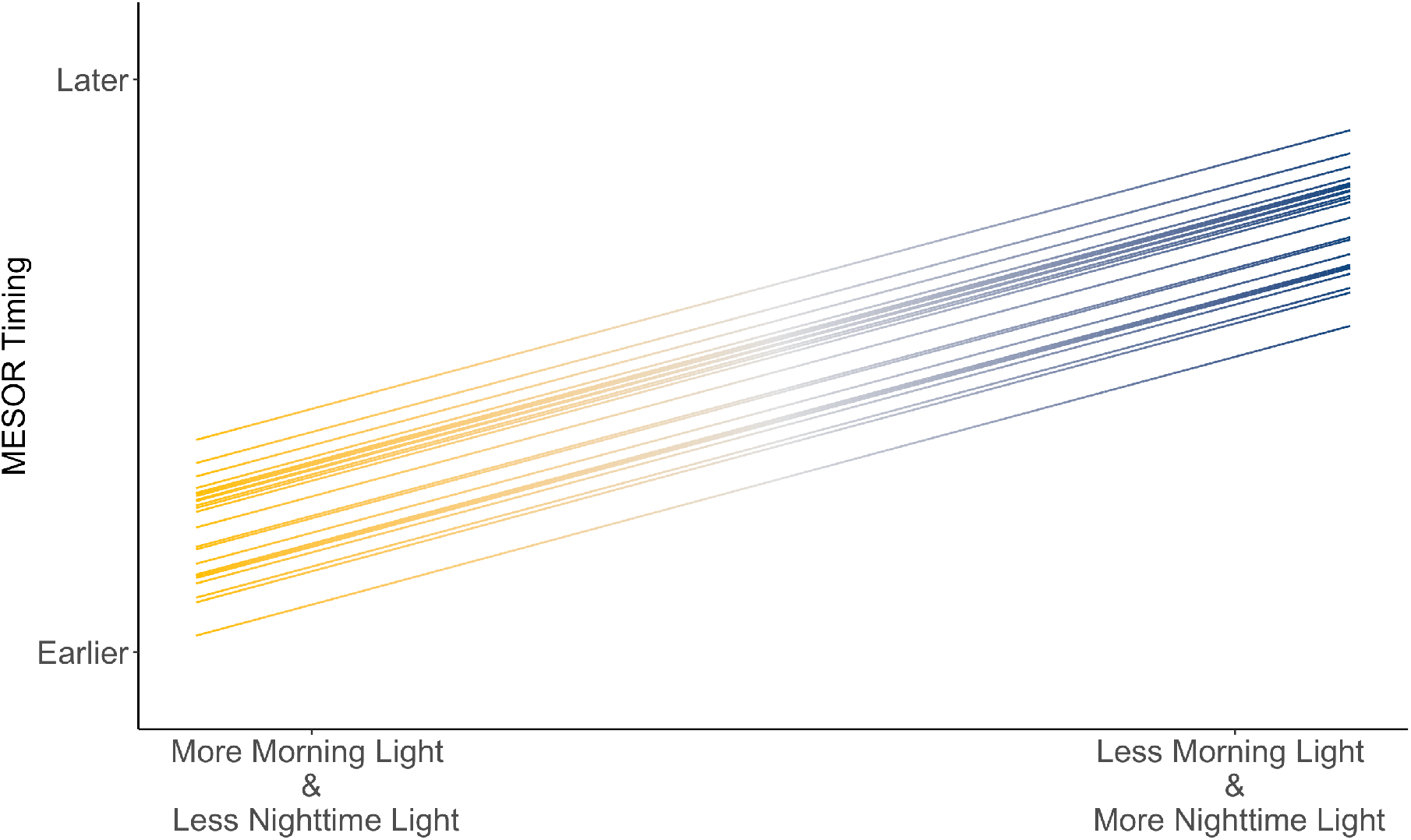
Conceptual Explanation of the effect of light exposure on MESOR timing. Projected change in MESOR timing at different levels of nighttime and morning light exposure, based on data outputs from the mixed model. Distance between parallel lines reflects difference in baseline MESOR timing intercept between individual participants.

## Discussion

This study characterized daily and seasonal light exposure and wrist temperature cycles in people living within their normal environment. Individual light exposure was significantly less than outdoor light across all four seasons. Low overall light exposure resulted from the use of artificial light during the day and what we assume to be little time spent outdoors, especially on weekdays. Electric light was also used at night, and this resulted in higher light exposure at night, relative to outdoor conditions. Individuals exhibited a wide range of light-at-night exposure, ranging from undetectable levels to those similar to daytime. Some individuals were so depauperate in daytime light, and enriched in nighttime light, that half of their total daily light exposure occurred at night. Due to our sensor’s inability to register light intensity values below 10 lux, we were unable to measure the effect of low intensity light-at-night and outdoor light pollution. Our evaluation of light exposure, and how it is partitioned among daytime and nighttime hours, supported our hypothesis that individuals living in urban environments dim out their days and light up their nights.

The results from our seasonal analysis lead us to conclude that there is minimal seasonal variation in the realized photoperiod experienced by individuals. We had hypothesized that people experience uniform light exposure throughout the year. We did, however, see some variation in light exposure, with a significant difference between summer and winter. However, this summer-winter difference in realized light exposure was one-fifth the magnitude of the same seasonal difference in the outdoor light sensors. The participants in our study live in relatively urban areas within a built environment that is heavily influenced by electric light. We cannot extrapolate our seasonal results to all urban environments, because some cities may be less dependent on electric light, facilitating more naturalistic seasonal cycles in light exposure. Furthermore, depending on economic/occupational and behavioral characteristics of a population, individuals living in other urban environments may have seasonal light exposure vastly different from that in New York City. Seasonal biology in humans is not well-understood, therefore, it is unknown whether the disconnect from natural light cycles we observed will have an impact on physiology and health.

Most importantly, this study revealed that differential light exposure, within the range seen in everyday life, can lead to regular shifts in circadian physiology within the general population. We identified alterations in circadian physiology in response to differential light exposure. Increased nighttime light shifted the morning MESOR timing later, while morning light shifted it earlier. There have been numerous elegant lab experiments demonstrating that drastic changes in light exposure (i.e., mimicking night shift and/or jet lag) can lead to circadian disruption [19]. To our knowledge, ours is one of the first studies to show there is an effect of differential light exposure on circadian rhythms in day-to-day life. It is important to note that individuals in our study kept relatively typical daily schedules, similar to that of a typical 9 AM-5 PM worker. Our results suggest, if we were to survey light exposure and circadian rhythms in a broader swath of the population, we may expect to find that circadian disruption is a more regular occurrence than has been recognized.

The large degree of variability in light exposure among individuals living in a similar geographic area, highlights the importance of personal light monitoring, as opposed to outdoor sensors and satellite data. Although light-at-night studies are highly represented in the chronobiology literature, we found that individuals experienced a high degree of variability in light exposure, not only at night, but across all hours of the day. This variability in light exposure may have broader implications for the generalizability of chronobiology studies conducted under strict experimental conditions. With the emerging focus on personalized medicine and the use of wearable devices to study behavior and health, we believe that the study of light exposure and circadian rhythms in real-time opens up new opportunities for individuals to harness their clock to improve health and wellbeing.

## Materials/Methods

### Recruitment/Data collection

This study was conducted under Columbia University IRB (Protocol Number AAAR7297 M00Y03). We recruited 23 adult participants for this study in summer 2018. Participants were recruited via flyers placed in Upper Manhattan, New York City (NYC) and Princeton, NJ. Inclusion into the study required the participants to state that they keep a relatively consistent 8-9 hour daily sleep schedule and did not identify as night owls. The majority of participants were from NYC (n = 19), with the mean age of participants being 32.2 years (sd = 8.33 years). We aimed to have a representative sample of individuals living in Northern Manhattan; 70% of the participants identified as women, 22% of the participants identified as Hispanic/Latino, 22% identified as Asian, 22% identified as Black/African American, 30% identified as White, and 4% identified as Other.

Participants were given light illuminance sensors (HOBO^®^ UA-002-08 Pendant Temperature/Light Data Logger) and wrist temperature sensors (iButton^®^ temperature loggers DS1922L/DS1922T). The light sensors had a lower limit of detection of 10 lux, which limited detection of low-intensity light exposure recorded. Refer to the supplemental information for a photo of the sensors. Each participant wore their sensors simultaneously for a full week during each of the four seasonal sampling sessions. The seasonal sampling sessions were held during weeks surrounding summer solstice 2018, autumn equinox 2018, winter solstice 2018, and spring equinox 2019. Loss of light/temperature sensors during observation periods and dropout between seasons lowered the effective sample size to 18 in the Summer, 16 in the Fall, 15 in the Winter, and 12 in the Spring.

### Light Exposure Characterization

Light exposure was measured in 5-minute intervals for each week-long seasonal sampling session, while outdoor light intensity was measured in 3-minute intervals over a two-week period each season. We aligned outdoor sampling sessions to match the timing of participant sampling. HOBO sensors were hung approx. 1.5 meters above ground facing north, typically on trees (refer to Supplemental Information for an image of the setup). At each outdoor sample location, one sensor was hung in a shaded location and another was hung in a well-lit location. Light illuminance, measured in lux, was log_10_ transformed for analyses. We analyzed data starting at 17:00 on the first day of sampling. Individual time series were categorized into observation days beginning at 17:00 and ending at 16:55 the following calendar day. Observation days were used when analyzing data over 24-hour periods. We created heatmaps to visualize changes in light illuminance over time, with each row containing each sequential light reading from within one observation day. Rows were organized to group together sequential observation days from the same light sensor within the same season.

We quantified light exposure as the area under the curve (AUC) for the log_10_lux time series using the trapezoidal rule. All AUC measurements are expressed with the unit log_10_lux-minutes. To study the seasonal variation in light exposure we measured (i) total daily light exposure, (ii) nighttime light exposure (i.e., sunset to 04:00), and (iii) daytime light exposure (i.e., 04:00 to approximate hour of sunset) for each observation day. We calculated this for both the participant data and the outdoor data for comparison. Tukey’s Honest Significant Difference test was used to compare means across seasons. To determine the effect of differential light exposure on circadian physiology, standardized nighttime (21:00 – 02:00) and morning (04:00 - 11:59) AUC was calculated for individual participants on each observation day. This was used as inputs for the linear mixed model described below. In order to quantify variability in light exposure at different times of day, the AUC was lastly calculated for four fixed-duration temporal windows: morning (05:00 – 11:00), afternoon (11:00 – 17:00), evening (17:00 – 23:00), and late night (23:00 – 05:00). The relative standard error of the AUC was calculated for each temporal window.

### Temperature Characterization

Wrist temperature was also measured in 5-minute intervals, synchronized with the light exposure measurements. Temperature readings outside of the normal biological range (< 29.5°C or > 38.5°C) were replaced with NAs, as we assumed these readings occurred when participants removed their device. To visualize global patterns in the relationship between light exposure and wrist temperature, time series matrices of light and temperature were identically gridded and treated as spatially-organized grids. Using these spatially-organized grids, we ran a Bivariate Local Moran’s I using queen contiguity and 8 orders of contiguity. The Moran’s I allowed us to identify significant clusters shared among the light and temperature matrices. Knowing that body temperature follows a periodic rhythm [20] with a period of approximately 24 hours, a periodic wave function was fit to each observation day of temperature readings by running a linear regression model with cos(2*pi*Time/24) and sin(2 * pi * Time/24) as predictor variables and temperature as the response variable. We then applied a goodness-of-fit measure to the periodic wave functions and only worked with those observation days where the average difference between predicted and observed wrist temperature was less than 1.11°C.

The fitted periodic wave functions were used to predict the timing of the MESOR (midline estimating statistic of rhythm) occurring between the peak and the trough following the daily maximum temperature within each observation day. A linear mixed model was then run with MESOR timing as the outcome, nighttime, morning, and afternoon light exposure as fixed effects, and individual participant number as the random effect (variable slope). The R package lme4 was used to fit the linear mixed model [21]. Accompanying p-values for the linear mixed model were calculated via Satterthwaite’s degrees of freedom method, using the R package lmerTest [22]. Additionally, multicollinearity among the input variables was tested for by calculating the variance inflation factor using the vif function from the R package usdm [23]. All data analysis was done in R version 3.6.2 [24]. Figures were generated using the R packages ggplot2 [25] and plotly [26].

## Supporting information

Light Study Supplement

## Data Availability

Data and analysis code have been made available at Dryad.

https://datadryad.org/stash/share/FzaYWTcL-gbiP7DXYx_sssrHYnWfPDkfk--FdYoWsS0

## Acknowledgements

Our research was funded by a pilot award from the NIEHS p30 Center (ES009089).

