## Supplementary material for "Characterizing the Modern Light Environment and its Influence on Circadian Rhythms": Light Study Supplement


**This PDF File includes:**

Supplemental Figures 1 to 4

Supplemental Tables 1 to 3

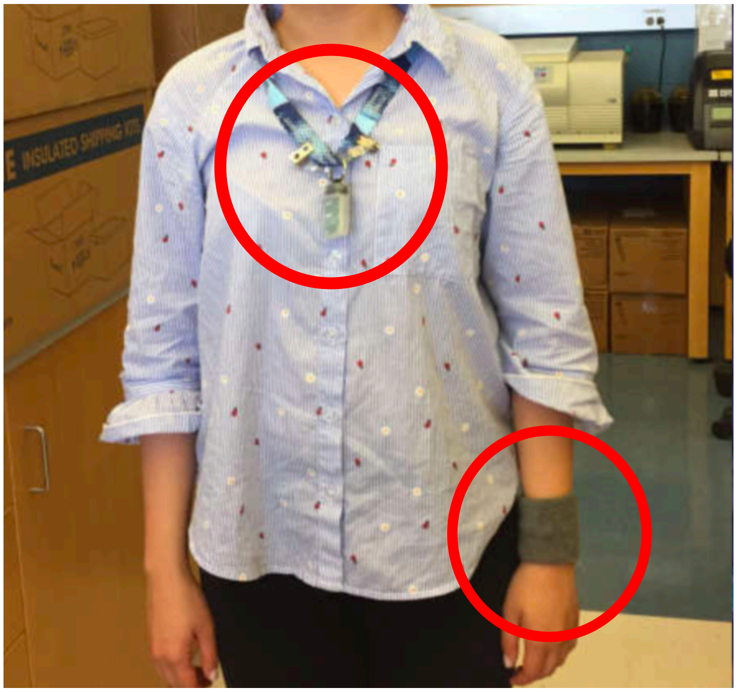

***Figure S1**: Example of the light sensor (worn around the neck) and wrist temperature sensor (worn around the wrist).*

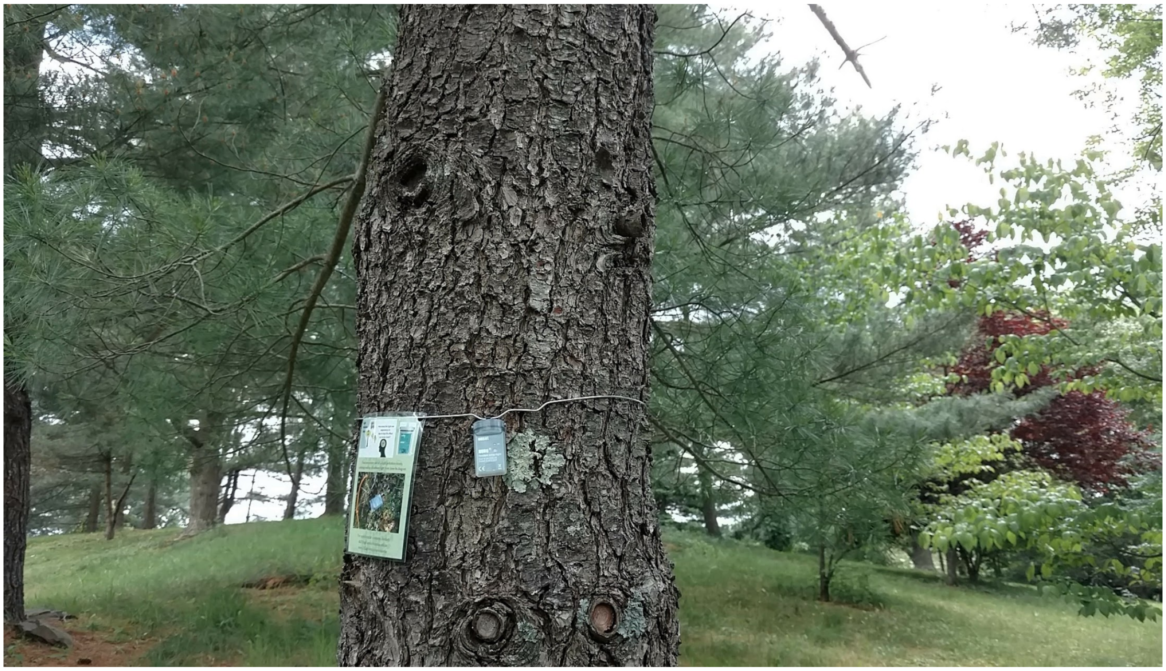

***Figure S2: Outdoor light sensor.*** *All outdoor light sensors were hung 1 meter off the ground facing North.*

| Comparison | Difference | Lower Est. | Upper Est. | p-value |
|---|---|---|---|---|
| **Outdoor Comparisons** | | | | |
| Spring-Fall | 716.55 | 281.99 | 1151.11 | *** |
| Summer-Fall | 1030.92 | 596.37 | 1465.48 | *** |
| Winter-Fall | -274.93 | -709.48 | 159.63 | ns |
| Summer-Spring | 314.37 | -120.19 | 748.92 | ns |
| Winter-Spring | -991.48 | -1426.04 | -556.93 | *** |
| Winter-Summer | -1305.85 | -1740.41 | -871.3 | *** |
| **Individual Comparisons** | | | | |
| Spring-Fall | -14.65 | -250.01 | 220.72 | ns |
| Summer-Fall | 115.46 | -89.08 | 319..99 | ns |
| Winter-Fall | -164.41 | -376.63 | 47.81 | ns |
| Summer-Spring | 130.1 | -100.3 | 360.51 | ns |
| Winter-Spring | -149.76 | -387.02 | 87.49 | ns |
| Winter-Summer | -279.87 | -486.58 | -73.16 | ** |
| **Individual vs Outdoor** | | | | |
| Fall | 972.31 | 631.01 | 1313.62 | *** |
| Winter | 861.79 | 519.18 | 1204.41 | *** |
| Spring | 1703.51 | 1346.1 | 2060.92 | *** |
| Summer | 1887.78 | 1549.87 | 2225.69 | *** |

***Table S1****: Tukey's Honest Significant Difference test results for the mean comparisons of total light exposure AUC summaries. Seasonal comparisons between Outdoor Sensors are shown in the Outdoor section. Seasonal comparisons between Individual Sensors are shown in the Individual section. Comparisons between Outdoor and Individual Sensors within the same season are shown in the Outdoor vs Individual section. (*** indicates <0.001, ** indicates <0.01, * indicates <0.05, ns indicates >0.05)*

**Random Effects**

| | Variance | Std. Dev. |
|---|---|---|
| Individual Number | 1.672 | 1.293 |
| Residual | 5.59 | 2.364 |

**Fixed Effects**

| | Estimate | Std. Error | df | t-value | Pr(>\|t\|) |
|---|---|---|---|---|---|
| (Intercept) | 1.65E+01 | 6.09E-01 | 1.08E+02 | 27.06 | 2.00E-16 |
| Morning Light | -4.00E-03 | 9.18E-04 | 2.59E+02 | -4.36 | 1.92E-05 |
| Nighttime Light | 2.80E-03 | 1.15E-03 | 2.48E+02 | 2.44 | 0.0155 |
| Afternoon Light | 8.28E-04 | 8.67E-04 | 2.62E+02 | 0.96 | 0.3401 |

**Scaled Residuals**

| Min | 1Q | Median | 3Q | Max |
|---|---|---|---|---|
| -2.792 | -0.571 | 0.023 | 0.463 | 3.266 |

***Table S2*: *Linear Mixed Model Full Summary Table*.** *Linear mixed model fit by REML, t-tests use Satterthwaite's method. Accompanying p-values for the fixed effects are under the "Pr(>|t|") column. Random effects allow for y-intercept to vary between individuals. All input variables had a variance inflation factor below 1.47, suggesting no multicollinearity among the variables.*

| | Individual Data | | Outdoor Data | |
|---|---|---|---|---|
| Season | Earliest Date | Latest Date | Earliest Date | Latest Date |
| Summer | June 21 | August 7 | July 2 | July 11 |
| Fall | September 6 | October 10 | October 3 | October 13 |
| Winter | January 3 | January 18 | January 5 | January 14 |
| Spring | March 25 | April 18 | April 5 | April 14 |

***Table S3: Observation periods for individual participants and outdoor sensors.***

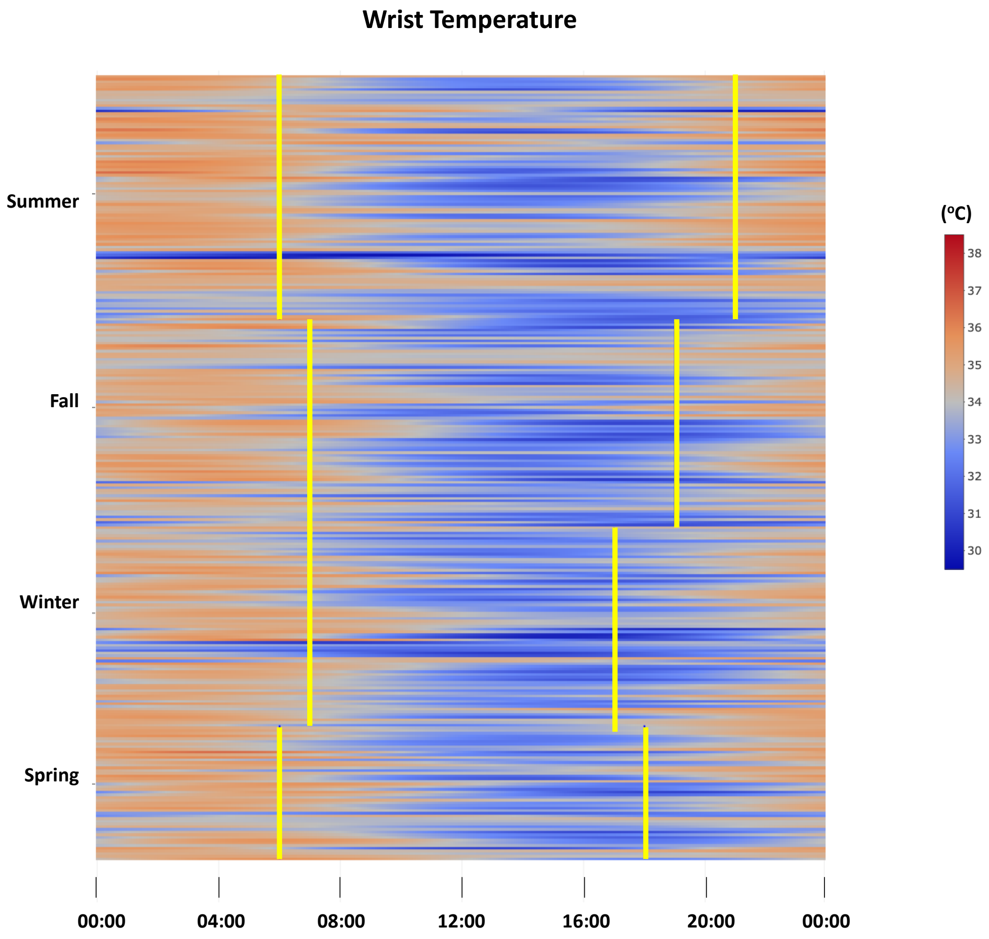


***Figure S3: Fitted periodic temperature data.*** *Predicted wrist temperature from the fitted periodic function.*

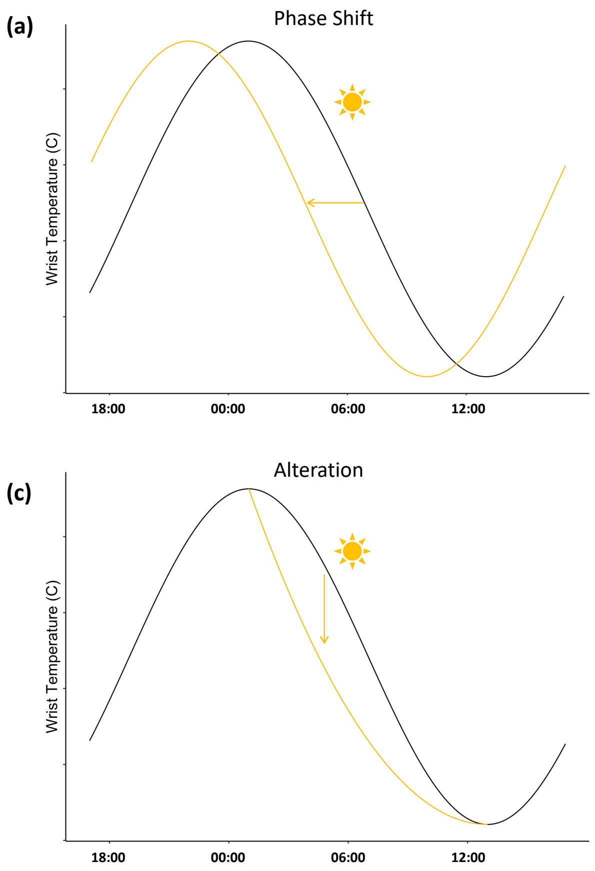


***Figure S4: Fitted periodic temperature data.*** *Predicted wrist temperature from the fitted periodic function. Proposed explanations for the effects of light exposure on wrist temperature rhythms. Normal temperature rhythms are shown in black. Shifted/altered temperature rhythms are shown in orange for the morning light and blue for the nighttime light.*
